# One year landscape of the inactivated virus vaccine triggered RBD-specific IgG, IgM and IgA responses against the Omicron variant

**DOI:** 10.1101/2022.01.02.22268647

**Authors:** Jun-biao Xue, Dan-yun Lai, He-wei Jiang, Jie Zhou, Sheng-ce Tao

## Abstract

We reported the first map of vaccine stimulated RBD-specific antibody responses (IgG, IgM and IgA) against the Omicron variant. We generated the map using a protein microarray, by analyzing longitudinal sera collected spanning one year from individuals immunized with 3 doses of an inactivated virus vaccine. The IgG response to RBD-Omicron is: 1/3-1/5 that of RBD-wild type; ∼6x higher for the booster dose *vs*. the 2^nd^ dose; and reaches the plateau in about two weeks after the booster dose, then drops ∼5x in another two weeks. Similar results were also obtained for IgM and IgA. Because of the high correlation between RBD-specific antibody response and the neutralization activity to authentic virus, we at least indirectly revealed the landscape of antibody protection against the Omicron variant throughout the vaccination stages. Our results strongly support the necessity of booster vaccination. However, post-booster vaccination may need to be considered.

---

The Omicron variant (B.1.1.529) possesses ∼32 mutations in its spike protein, and at least 15 amino acid changes in the receptor binding domain (RBD)^1^. The Omicron variant is worsening the COVID-19 pandemic largely due to the significant escape from the neutralization antibodies^2^. To combat Omicron variant, one of the most effective ways is vaccination, especially the booster vaccination. In China and some other countries, the inactivated virus vaccine is dominant^3^. And because of the huge population, the vaccination campaign is carried out sequentially in China, start from at-risk groups and big cities, then to other people and other places. Thus, across the country, many people are at different vaccination stages, *i. e*., unvaccinated, the 1^st^ dose, the 2^nd^ dose and the 3^rd^ (booster) dose. It is of great interest to know what level of protection can the inactivated virus vaccine provide against the Omicron variant for the people at varied vaccination stages. Which will guide us to optimize or design the long-term vaccination program.

## Results

### The Omicron RBD protein microarray

It is well known that RBD-specific IgG response is highly correlated to the neutralization activity of authentic SARS-CoV-2 virus^4^. For fast and systematical evaluation of the neutralization activity against the Omicron variant, we constructed a protein microarray by including the wild-type and the Omicron RBD, denoted as RBD-WT and RBD-Omicron (**Fig. 1a and 1b**).

**Fig 1.**
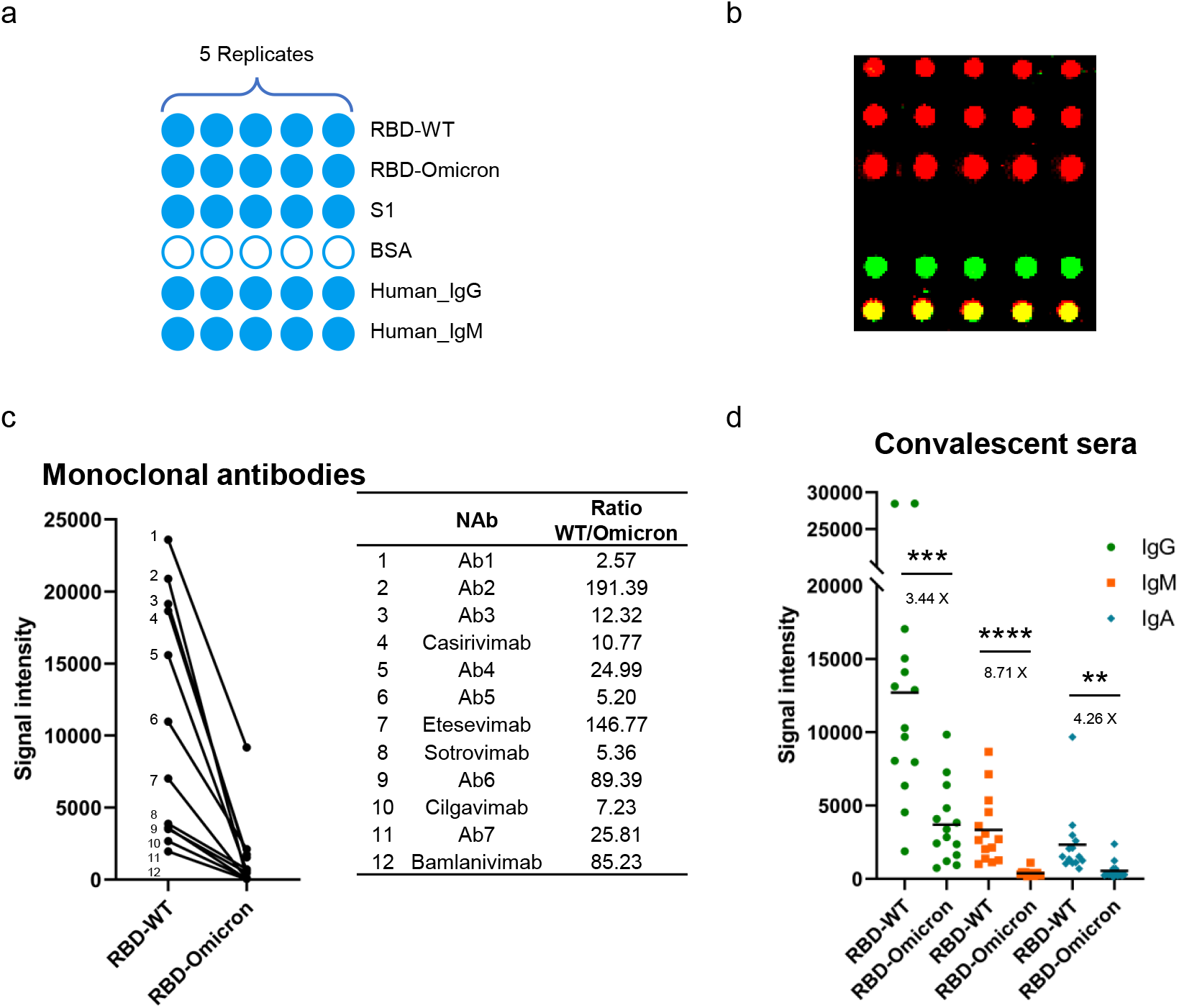
The Omicron RBD protein microarray. **a**, The layout of the protein microarray; **b**, Quality inspection of the protein microarray which was probed with anti-6xHis, anti-Human_IgG and anti-Human_IgM antibodies; **c-d**, The bindings of RBD-specific monoclonal antibodies on the microarray (**c**) and 14 COVID-19 convalescent sera (**d**) against RBD-WT (wild type) and RBD-Omicron. The black horizontal lines indicate the average signal intensity. Numbers in black (**d**) denote fold-change in signal intensity between RBD-WT and RBD-Omicron.

To assure the applicability of the protein microarray, we tested 12 SARS-CoV-2 RBD specific monoclonal antibodies on the microarray (**Fig. 1c**). Lower bindings to RBD-Omicron were observed for all the antibodies, with the ratio RBD-WT/ RBD-Omicron ranges from 2.57 to 191.39. This indicates that most of these antibodies may significantly lose their neutralization activity to Omicron variant. Among these antibodies, Casirivimab, Etesevimab,Bamlanivimab,Cilgavimab and Sotrovimab were also tested in a recent study^5^. Specifically, for Casirivimab, Etesevimab and Bamlanivimab, we observed 10.77x, 146.77x and 85.23x lower binding to Omicron-RBD, while completely abolished neutralization activities were reported for all these three antibodies. For Cilgavimab and Sotrovimab, we observed 7.23x and 5.36x lower binding to Omicron-RBD, while 20x and 3x lower neutralization activities were reported. Thus, our data are generally consistent to that of the ELISA assay and the authentic Omicron variant neutralization assay, suggest that the protein microarray could serve as alternative but more convenient approach for fast assessment of the neutralization activities of samples to the authentic Omicron variant. In addition, our results support the expectation that many of the current mAbs may largely or completely lose their neutralization activities to the Omicron variant.

To further confirm the applicability of the protein microarray, 14 convalescent sera (**Table S1**-**cohort 1**) were analyzed following an established protocol ^6,7^, besides IgG, IgM and IgA responses were also compared between RBD-WT and RBD-Omicron. Significantly lower signals were observed for all these 3 types of antibodies against RBD-Omicron, specifically, 3.44x, 8.71 and 4.26x for IgG, IgM and IgA, respectively. The IgG signal is consistent to recent studies^8^. Furthermore, to our knowledge, we are the first to report the decrease of the IgM and IgA responses to the Omicron variant. Our results suggest the protection from the previous wild-type virus infection to the Omicron variant need to be strengthened through an appropriate way.

### RBD-specific IgG responses of longitudinal sera from individuals immunized with inactivated virus vaccine BBIBP-CorV

Throughout 2021, we have collected sera from a cohort of 13 individuals who have been fully vaccinated (the 1^st^ dose, the 2^nd^ dose and the booster dose) with the inactivated virus vaccine BBIBP-CorV at 16 time points (**Fig. 2a, Table S1**-**cohort 2**).

**Fig. 2.**
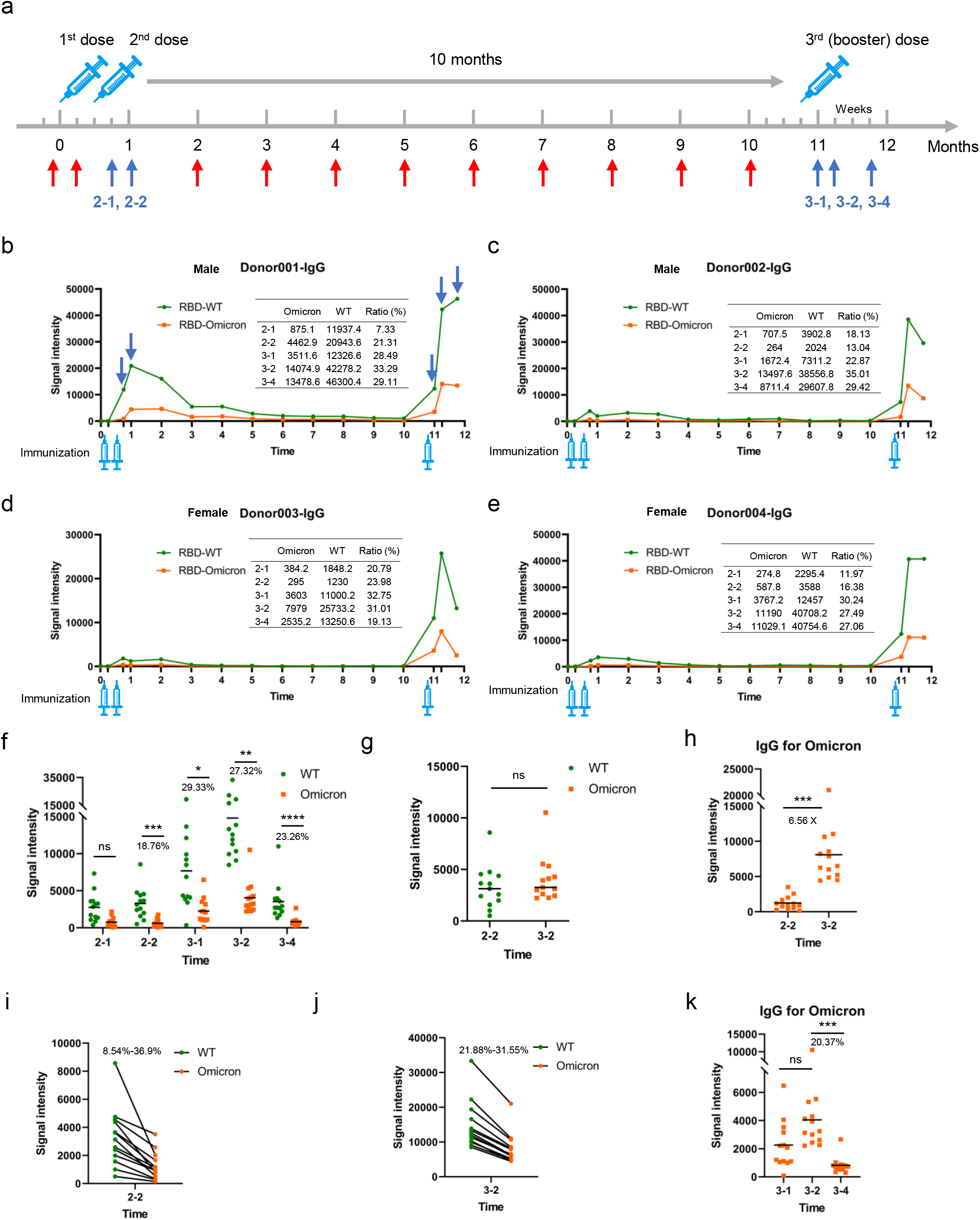
RBD-specific IgG responses of longitudinal sera collected from vaccinated individuals. **a**, Scheme of immunization and sampling of the longitudinal cohort, vertical arrows indicate the 16 time points for collecting sera; **b-e**, The trends of RBD-specific IgG responses during one year for 4 individuals that were immunized with 3 doses of inactivated virus vaccine; **f-k**, IgG responses against RBD-WT and RBD-Omicron at 5 time points that with relatively higher antibody levels, *i. e*., 2-1 and 2-2, represent one and 2 weeks after the 1^st^ dose, respectively; 3-1, 3-2 and 3-4 represent 1, 2 and 4 weeks after the 3^rd^ (booster) dose, respectively. The overall differences at the 5 time points between RBD-WT and RBD-Omicron (**f**). The IgG responses against RBD-Omicron at 3-2 and against RBD-WT at 2-2 (**g**). The IgG responses against RBD-Omicron at 2-2 and 3-2 (**h**). The IgG responses between RBD-WT and RBD-Omicron at 2-2 (**i**) and 3-2 (**j**). The trends of Omicron-RBD specific IgG responses after the 3^rd^ dose (**k**).

To learn what IgG level can the inactivated virus vaccine provide against the Omicron variant for the people at varied vaccination stages, serum samples of cohort2 were subjected for protein microarray analysis. We first analyzed the full set of samples (16 time points) of four individuals (Donor001-Donor004) (**Fig. 2b-2e**). From the 1^st^ to the 16^th^ time points, similar trends of IgG responses were observed for all the four individuals. Take Donor001 as an example (**Fig. 2b**), the trends of IgG responses were similar between RBD-WT and RBD-Omicron, specifically, the IgG response was barely detectable 1 week after the 1^st^ dose, while obvious response was observed 1 week after the 2^nd^ dose, and the response reached the plateau 2 weeks after the 2^nd^ dose, then gradually descended till the booster dose. After the booster dose, the IgG response was sharply increased and reached the plateau around 2-4 weeks, overall, the IgG response of the booster dose is much higher than that of the 2^nd^ dose. For all the 16 tome points, the IgG responses against RBD-Omicron are significantly lower than that of RBD-WT.

To further illustrate the difference of IgG responses, IgG responses against RBD-WT and RBD-Omicron at 5 time points that with relatively higher antibody levels were analyzed for all the 13 individuals (**Table S1**-**cohort 2)**, *i. e*., 2-1 and 2-2, represent 1 and 2 weeks after the 2^nd^ dose, respectively; 3-1, 3-2 and 3-4 represent 1, 2 and 4 weeks after the 3^rd^ dose, respectively (**Fig. 2f**). Except 2-1, for the other 4 time points, the averaged IgG responses to Omicron-RBD ranged from 18.76% to 29.33% to that of RBD-WT, and the differences were statistically significant.

It is of great interest to see the protection that the booster vaccination can provide for the Omicron variant. We compared the highest IgG responses to RBD-Omicron after the booster dose, *i. e*., 3-2 to the highest responses to RBD-WT after the 2^nd^ dose, *i. e*., 2-2. Comparable IgG responses were observed (**Fig. 2g**). We also noticed that RBD-Omicron IgG response at 3-2 is 6.56x to that of 2-2 (**Fig. 2h**). These results indicates that, to combat the Omicron variant, the booster dose is necessary, and may could provide comparable protection to the Omicron variant as that of the 2^nd^ dose to the wild type strain.

According to full time points analysis of the four individuals (**Fig. 2b-2e)**, the RBD-Omicron IgG were lower than that of RBD-WT at all the time points. To check whether this is also the case, we picked 2-2 and 3-2, and performed side-by-side analysis. Our expectation was confirmed (**Fig. 2i, 2j**).

It is known that the IgG responses to the wild-type strain gradually decrease after the plateau of the 2^nd^ dose, will it be the same for the Omicron variant after the booster dose? To answer this, we compared the RBD-Omicron IgG responses among 3-1, 3-2 and 3-4. The average signal is only 20.37% left for 3-4 as compared to that of 3-2 (**Fig. 2k**). This result indicates an unexpected sharp decrease of protection against Omicron variant in only two weeks.

### RBD-specific IgM responses of longitudinal sera from individuals immunized with BBIBP-CorV

The IgM responses are also of interest. To learn what IgM level can the inactivated virus vaccine provide against Omicron variant for the people at varied vaccination stages, serum samples of cohort2 were subjected for protein microarray analysis. We first analyzed the full set of samples (16 time points) of four individuals (Donor001-Donor004) (**Fig. 3a-3d**). From the 1^st^ to the 16^th^ time points, similar trends of IgM responses were observed for all the four individuals. Take Donor001 as an example (**Fig. 3a**), the trends of IgM responses were similar between RBD-WT and RBD-Omicron, specifically, the IgM response was undetectable 1 week after the 1^st^ dose, while it was obvious 1 week after the 2^nd^ dose, and the response reached the plateau 2 weeks after the 2^nd^ dose, then descended slowly till the booster dose. After the booster dose, the IgM response was increased and reached the plateau around 2-4 weeks, overall, the IgM response after the booster dose is slightly higher than that of the 2^nd^ dose. For all the 16 tome points, the IgM responses against the RBD-Omicron are lower than that of the RBD-WT.

**Fig. 3.**
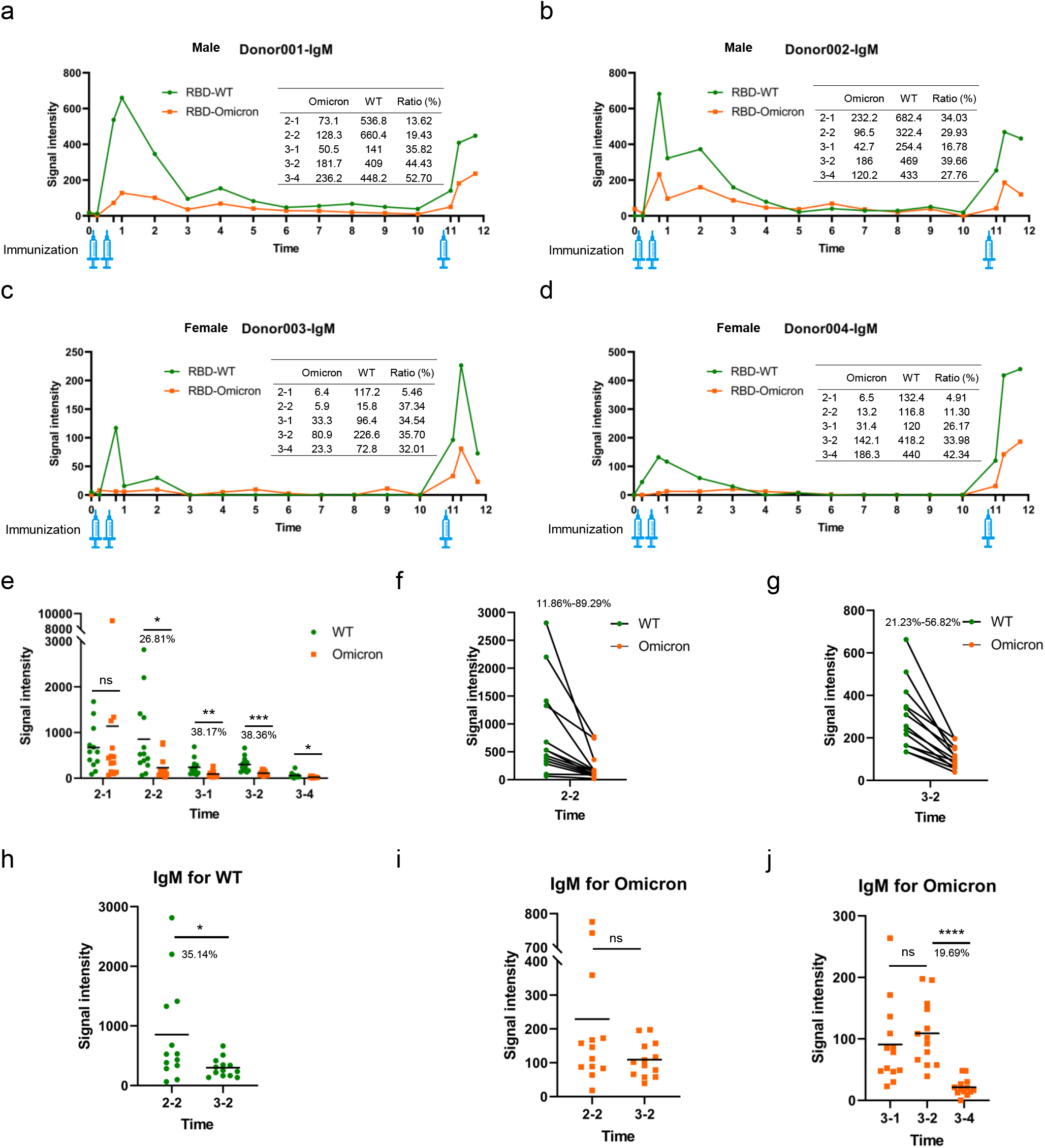
RBD-specific IgM responses of longitudinal sera collected from vaccinated individuals. **a-d**, The trends of RBD-specific IgM responses during one year for 4 individuals that were immunized with 3 doses of inactivated virus vaccine; **e-j**, IgM responses against RBD-WT and RBD-Omicron at the 5 time points. The overall differences at the 5 time points between RBD-WT and RBD-Omicron (**e**). The IgM responses between RBD-WT and RBD-Omicron at 2-2 (**f**) and 3-2 (**g**). The IgM responses at 2-2 and 3-2 against RBD-WT (**h**) and RBD-Omicron (**i**), respectively. The trends of Omicron-RBD specific IgG responses after the 3^rd^ dose (**j**).

To further illustrate the difference of IgM responses, IgM responses against RBD-WT and RBD-Omicron at 5 time points that with relatively higher antibody levels were analyzed for all the 13 individuals (**Table S1**-**cohort 2)** (**Fig. 3e**). Except 2-1, for the other 4 time points, the averaged IgM responses to the Omicron-RBD ranged from 26.81% to 38.36% to that of the RBD-WT, and the differences were statistically significant. According to full time points analysis of the four individuals (**Fig. 3a-3d)**, the RBD-Omicron IgM responses were lower than that of the RBD-WT at all the time points. To check whether this is also the case for other individuals, we picked 2-2 and 3-2, and performed side-by-side analysis. Our expectation was confirmed (**Fig. 3f, 3g**). To further illustrate the results of **Fig. 3e**, we compared the IgM responses to RBD-WT at 2-2 and 3-2 (**Fig. 3h**), RBD-Omicron at 2-2 and 3-2 (**Fig. 3i**). The IgM responses were averagely lower at 3-2 for both RBD-WT and RBD-Omicron.

It is known that the IgG responses to the wild-type strain gradually decrease after the plateau of the 2^nd^ dose, will it be the same for IgM responses to the Omicron variant after the booster dose? To address this, we compared the RBD-Omicron IgM responses among 3-1, 3-2 and 3-4. The average signal is only 19.69% left for 3-4 as compared to that of 3-2 (**Fig. 3j**). This result indicates a sharp decrease of IgM against Omicron variant in two weeks, that is explainable.

### RBD-specific IgA responses of longitudinal sera from individuals immunized with BBIBP-CorV

Mucosal immunity is a key component of our immune system to fight SARS-CoV-2, and IgA plays a major role in mucosal immunity^9,10^.

To learn what IgA level can the inactivated virus vaccine provide against Omicron variant for the people at varied vaccination stages, serum samples of cohort2 were subjected for protein microarray analysis. We first analyzed the full set of samples (16 time points) of four individuals (Donor001-Donor004) (**Fig. 4a-4d**). From the 1^st^ to the 16^th^ time points, similar trends of IgA responses were observed for all the four individuals. Take Donor001 as an example (**Fig. 4a**), the trends of IgA responses were similar between RBD-WT and RBD-Omicron, specifically, the IgA response was barely detectable at 1 week after the 1^st^ dose, while obvious response was observed 1 week after the 2^nd^ dose, and the response reached the plateau 2 weeks after the 2^nd^ dose, then gradually descended till the booster dose. After the booster dose, the IgA response was sharply increased and reached the plateau around 2 weeks, overall, the IgA response of the booster dose is higher than that of the 2^nd^ dose. For all the 16 tome points, the IgA responses against the RBD-Omicron are significantly lower than that of the RBD-WT.

**Fig. 4.**
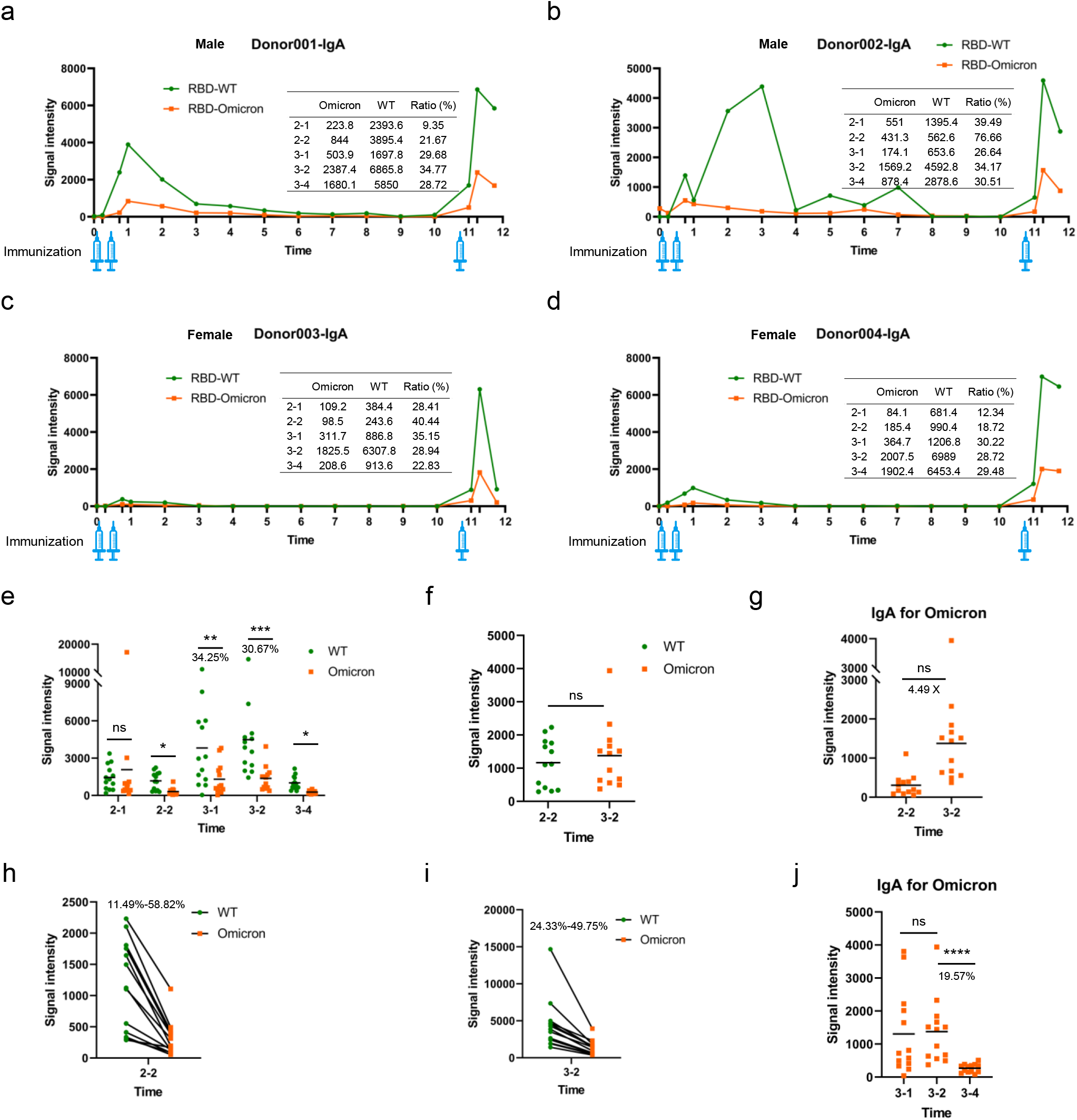
RBD-specific IgA responses of longitudinal sera collected from vaccinated individuals. **a-d**, The trends of RBD-specific IgA responses during one year for 4 individuals that were immunized with 3 doses of inactivated virus vaccine; **e-j**, IgA responses against RBD-WT and RBD-Omicron at the 5 time points that with relatively higher antibody levels. The overall differences at the 5 time points between RBD-WT and RBD-Omicron (**e**). The IgA responses against RBD-Omicron at 3-2 and against RBD-WT at 2-2 (**f**). The IgA responses against Omicron-RBD at 2-2 and 3-2 (**g**). The IgA responses against RBD-WT and RBD-Omicron at 2-2 (**h**) and 3-2 (**i**). The trends of Omicron-RBD specific IgA responses after the 3^rd^ dose (**j**).

To further illustrate the difference of IgA responses between the RBD-Omicron and RBD-WT, IgA responses against RBD-WT and RBD-Omicron at 5 time points that with relatively higher antibody levels were analyzed for all the 13 individuals (**Table S1**-**cohort 2)** (**Fig. 4e**). Except 2-1, for the other 4 time points, the averaged IgG responses to the Omicron-RBD were ∼30% to that of the RBD-WT, and the differences were statistically significant.

If is of great interest to see the mucosal immunity that the booster vaccination can provide for the Omicron variant. We compared the highest IgA responses to RBD-Omicron after the booster dose, *i. e*., 3-2 to the highest responses to RBD-WT after the 2^nd^ dose, *i. e*., 2-2. Comparable IgA responses were observed (**Fig. 4f**). We also noticed that RBD-Omicron IgA response at 3-2 is 4.49x to that of 2-2 (**Fig. 4g**). These results suggests that, to combat the Omicron variant, the booster dose is necessary, and may could provide comparable protection to the Omicron variant as that of the 2^nd^ dose to the wild type strain.

According to full time points analysis of the four individuals (**Fig. 4a-4d)**, the RBD-Omicron IgA were lower than that of the RBD-WT at all the time points. To check whether this is also the case for the whole cohort, we picked 2-2 and 3-2, and performed side-by-side analysis. Our expectation was confirmed (**Fig. 4h, 4i**).

It is known that the IgG responses to the wild-type strain gradually decrease after the plateau of the 2^nd^ dose, will it be the same for the IgA responses to the Omicron variant after the booster dose? To address this, we compared the RBD-Omicron IgA responses among 3-1, 3-2 and 3-4. The average signal is only 19.57% left for 3-4 as compared to that of 3-2 (**Fig. 4j**). This result indicates an unexpected sharp decrease of mucosal immunity from Omicron variant in only two weeks.

## Discussion

Our results clearly demonstrated that Omicron variant could escape the binding of a variety of neutralization antibodies, from slightly lower to almost abolished^2^. For convalescent sera, lower bindings to Omicron RBD indicate when facing Omicron variant, patient who recovered from previous infection is still in danger. For people who were vaccinated with the inactivated virus vaccine, significantly lower antibody bindings to the Omicron *vs*. wild type RBD during the whole process of vaccination were observed, which suggest the protection of the current vaccine for combating Omicron variant is reduced, but still effective^8,11^.

Our study has some limitations. The accessibility to biosafety facility is very limited, thus we haven’t confirmed our results on authentic wild type virus and Omicron variant. However, because of the high correlation between RBD binding and neutralization activity of authentic virus, the flexible microarray platform is suitable for fast evaluation. The cohort is small, however, to reduce the person-to-person variation, longitudinal samples collected throughout 1 year were applied. Our findings could be strengthened by a bigger cohort. Because of the availability, we only analyzed samples collected from people vaccinated with the inactivated virus vaccine BBIBP-CorV. It will also be interest to analyze longitudinal samples of other type of vaccines.

The major findings of this study are : 1) The 1^st^ year-long landscape of RBD-specific IgG, IgM and IgA responses against Omicron variant; 2) The RBD-specific IgG response of the Omicron variant is ∼1/3-1/5 of the wild type strain for both the 2^nd^ and the booster doses of the inactivated virus vaccination; 3) RBD-specific IgG response of the Omicron variant after the booster vaccination is averagely 6.56x higher than that of the second dose; 4) The RBD-specific IgG response of the Omicron variant after the booster vaccination reaches the plateau around 2 weeks and then drops to 20.37% after another two weeks. Based on these findings, we now have an overall picture about what level of protection may the inactivated virus vaccine provides against the Omicron variant for the people at varied vaccination stages. Our results support that the necessity of the booster vaccination. And more importantly, the vaccination plan after the booster vaccination may have to be considered.

## Methods

### Protein microarray fabrication

The RBD proteins (Acro Biosystem, Beijing, China) and S1 protein (Abclonal, Wuhan, China) of SARS-CoV-2, along with the negative (BSA) and positive controls (Human IgG and IgM), were printed in five replicates on PATH substrate slide (Grace Bio-Labs, Oregon, USA) to generate identical arrays in a 2 × 7 subarray format using Super Marathon printer (Arrayjet, UK). The microarrays were stored at -80°C until use.

### Patients and samples

All the RBD-specific mAbs were provided by Sanyou Bioparmaceuticals (Shanghai, China). A total of 14 COVID-19 convalescent patients and 13 participants of vaccination were enrolled in this study. The patients were hospitalized and received treatment in Foshan Forth hospital, the serum samples were collected on the day of hospital discharge (Cohort 1 in **Table 1**). A total of 13 participants of vaccination were recruited from the beginning of 2021 (Cohort 2 in **Table 1**), the participants were vaccinated with the inactivated virus vaccine BBIBP-CorV, and longitudinal serum samples were collected at 16 time points during one year, the samples included unvaccinated, the 1^st^ dose, the 2^nd^ dose and the 3^rd^ (booster) dose (**Figure 2A**).

### Microarray-based serum and antibody analysis

A 14-chamber rubber gasket was mounted onto each slide to create individual chambers for the 14 identical subarrays. The microarray was used for serum profiling as described previously with minor modifications^12^. Briefly, the arrays were brought from -80°C to - 20°C and room temperature for gradient rewarming and then incubated in blocking buffer (3% BSA in 1×PBS buffer with 0.1% Tween 20) for 3 h. A total of 200 μL of diluted sera was incubated with each subarray overnight at 4°C, monoclonal antibodies was incubated 2h at RT. The sera were diluted at 1:200. The antibody was diluted to a final concentration of 10 μg/mL for incubation. The microarrays were washed with 1×PBST and the bound antibodies were detected by incubating with Cy3-conjugated goat anti-human IgG and Alexa Fluor 647-conjugated donkey anti-human IgM (Jackson ImmunoResearch, PA, USA), which were diluted for 1:1,000 in 1×PBST. The incubation was carried out at room temperature for 1 h. The microarrays were then washed with 1×PBST and dried by centrifugation at room temperature. The microarrays were scanned by LuxScan 10K-A (CapitalBio Corporation, Beijing, China) with the parameters set as 100% laser power/ PMT 550, 100% laser power/ PMT 500 for IgM and IgG (Cy5 and Cy3 channel), respectively. For IgA (Fluorescein channel), the microarrays were scanned by GenePix 4200A (Molecular Devices, CA, USA) with the parameters set as 100% laser power/PMT 400. The fluorescent intensity was extracted by GenePix Pro 6.0 software (Molecular Devices, CA, USA).

### Ethical approval

The study was approved by the Institutional Ethics Review Committee of Foshan Fourth Hospital, Foshan, China (ref. no. 202005) and the Ethics Commission of Shanghai Jiao Tong University (ref. no. B2021120I). The written informed consent was obtained from each participant. Participants of vaccination have been informed the sera collection after booster vaccination and the written informed consent was obtained from each participant.

### Data analysis of protein microarray

For each spot, signal intensity was defined as the median_foreground subtracted by the median_background. The signal intensities of the five replicates spots for each peptide or protein were averaged. IgG, IgM and IgA data were analyzed separately. *P-values* for statistical analysis were calculated by unpaired two-way *t*-test. All diagram and statistical analyses were carried out using GraphPad Prism 8 software.

### Data availability

The SARS-CoV-2 RBD protein microarray data are deposited on Protein Microarray Database under the accession number PMDE260. Additional data related to this paper may be requested from the corresponding author.

## Acknowledgements

We thank the participants for their willingness and great support to be part of the longitudinal SARS-CoV-2 serum profiling and vaccine related study. It will not be possible without their great contribution. We thank Mr. Chengliang Zhang and Mrs. Dongxue Chai for their help on sample collection. We thank Dr. Guojun Lang for providing the antibodies. We thank Dr. Daniel M. Czajkowsky for critical reading and editing. This work was partially supported by the National Key Research and Development Program of China Grant (No.2016YFA0500600), National Natural Science Foundation of China (No. 31970130).

## Author contributions

Conceptualization: SCT. Methodology: JBX and DYL. Convalescent sample cohort design: JZ. Key materials: HWJ. Writing: SCT, JBX and DYL.

## Additional information statement

Correspondence and requests for materials should be addressed to Sheng-ce Tao.

## Figure legends

**Table S1.** Information of samples used in this study.

|  |  | Cohort 1 |  | Cohort 2 |
| --- | --- | --- | --- | --- |
| Group |  | Convalescent |  | Vaccination |
| Numbers |  | 14 |  | 13 |
| Age |  | 44.7±15.5 |  | 40.2±7.9 |
| Gender | Male | 7 |  | 11 |
|  | Female | 7 |  | 2 |
| Severity | Severe | 0 |  |  |
|  | non-severe | 14 |  | - |
|  | Death | 0 |  |  |
| Days after onset |  | 25.6±4.9 |  | - |
| Source |  | Foshan 4th Hospital, Guangdong |  | Shanghai Jiao Tong University, Shanghai |

